# Metabolic variation reflects dietary intake in a multi-ethnic Asian population

**DOI:** 10.1101/2023.12.04.23299350

**Authors:** Dorrain Yanwen Low, Theresia Handayani Mina, Nilanjana Sadhu, Kari E Wong, Pritesh Rajesh Jain, Rinkoo Dalan, Hong Kiat Ng, Wubin Xie, Benjamin Lam, Darwin Tay, Xiaoyan Wang, Yik Weng Yew, James Best, Rangaprasad Sarangarajan, Paul Elliott, Elio Riboli, Jimmy Lee, Eng Sing Lee, Joanne Ngeow, Patricia A Sheridan, Gregory A Michelotti, Marie Loh, John Chambers

**Affiliations:** Lee Kong Chian School of Medicine, Nanyang Technological University, Singapore; Department of Endocrinology, Tan Tock Seng Hospital, Singapore; Integrated Care for Obesity & Diabetes, Khoo Teck Puat Hospital, Singapore; National Skin Centre, Singapore; Department of Epidemiology and Biostatistics, Imperial College London, London, UK; Department of Psychosis, Institute of Mental Health, Singapore; Clinical Research Unit, National Healthcare Group Polyclinic, Nexus@one-north, Singapore; Division of Medical Oncology, National Cancer Centre, Singapore; Metabolon, Morrisville, North Carolina, USA

## Abstract

Dietary biomarkers reflecting habitual diet are explored largely in European and American populations. However, the “food metabolome” is highly complex, with its composition varying to region and culture. Here, by assessing 1,055 plasma metabolites and 169 foods/beverages in 8,391 comprehensively phenotyped individuals from the multi-ethnic Asian HELIOS cohort (69% Chinese, 12% Malay, 19% South Asian), we report novel observations for ethnic-relevant and common foods. Using machine-learning feature selection approach, we developed dietary multi-biomarker panels (3-39 metabolites each) for key foods and beverages in respective training sets. These panels comprised distinct and shared metabolite networks, and captured variances in intake prediction models in test sets better than single biomarkers. Composite metabolite scores, derived from the biomarker panels, associated significantly and more strongly with clinical phenotypes (HOMA-IR, type 2 diabetes, BMI, fat mass index, carotid intima-media thickness and hypertension), compared to self-reported intakes. Lastly, in 235 individuals that returned for a repeat visit (averaged 322 days apart), diet-metabolite relationships were robust over time, with predicted intakes, derived from biomarker panels and metabolite scores, showing better reproducibility than self-reported intakes. Altogether, our findings show new insights into multi-ethnic diet-related metabolic variations and new opportunity to link exposure to health outcomes in Asian populations.

## Main

Non-communicable diseases (NCDs) including obesity, diabetes and cardiovascular diseases are leading global causes of death and disability. Although there are established genetic and environmental contributors to NCD risk, modifiable lifestyle determinants such as diet, play a major role^1^. The marked rise in NCDs has a causal link to global dietary habits becoming increasingly urbanised, being characterised by higher intakes of processed foods, refined grains and saturated fat, and concomitant lack of fibre-rich foods^2^. This connection emphasises the importance of accurate assessment of dietary intakes, and their relationship to health, to underpin nutritional recommendations and policy formulations.

Assessment of dietary intakes in epidemiological or interventional studies typically is a 2-step process involving self-report and subsequent estimation of daily/weekly intakes using food and/or nutrient composition databases. Limitations of self-reporting tools are well-documented including poor reproducibility, recall bias, misjudgement in portion sizes/frequency, and restricted list of foods and/or beverages^3,4^, which are compounded by variability in composition databases. These limitations may attenuate diet-disease relationships or lead to spurious findings.

Profiling our “food metabolome” using untargeted approaches has emerged as a valuable strategy in discovery of dietary biomarkers^5^ as objective measures of habitual dietary exposures. This approach has created awareness of the untracked diversity of compounds in foods that remains largely invisible to epidemiological or hypothesis-driven nutrition studies^6^. These diet-derived metabolites reflect true concentrations in biofluids (blood, urine) and could potentially overcome limitations of current dietary instruments, while also accounting for determinants of interindividual variability in metabolism including genetic background and gut-microbial composition^7,8^. Dietary biomarkers have been identified and validated for coffee^9^, orange^10^, banana^11^ and red meat^12^ in European populations but are lacking for other foods, foods consumed as part of complex dietary patterns and/or relevant to populations in other regions. For example, different biomarkers of coffee intake, rather than a unique biomarker, were identified across four European populations^13^, corresponding to how coffee was prepared in each country. Considering the regional and cultural diversity of diet across the world, biomarkers relevant to Asian populations are lacking, compared to common/Westernised foods.

In this study, we aim to i) identify relevant dietary biomarker panels reflecting food and beverage intakes within a multi-ethnic Asian population, and ii) develop metabolite scores to improve accuracy of assessing clinical associations with diet.

## Results

### Study design

Baseline dietary, clinical and biological measurements were collected from 10,004 free-living adults from the Singapore population. Longitudinal data were recollected in a random subset of individuals (n=235) on average one year after their baseline visit (range 58-1025 days), to enable assessment of reproducibility. The study design is illustrated in Fig. 1. Further details are provided in Methods and Extended Data Table 1.

**Fig. 1.**
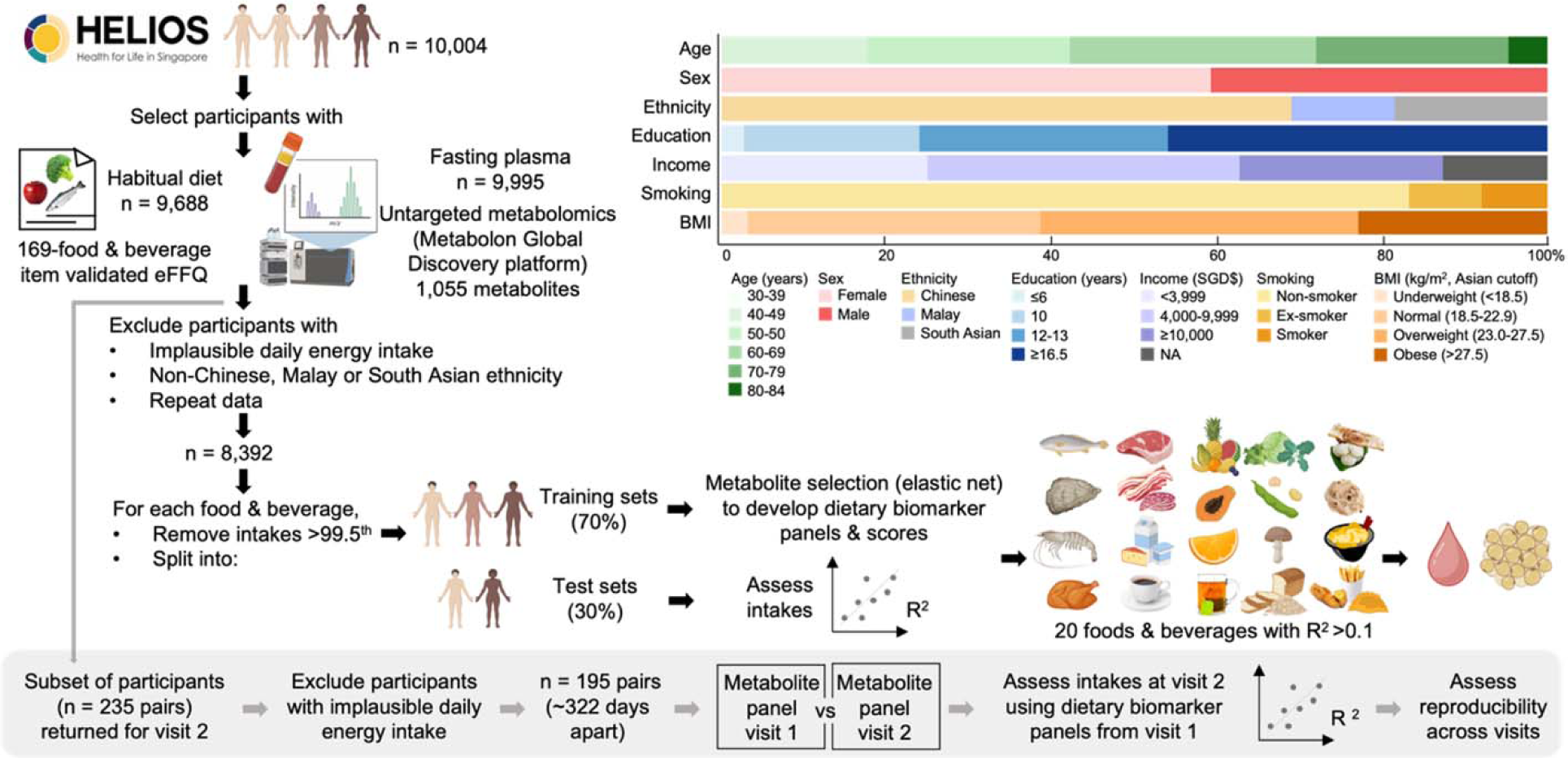
Overview of study design. The HELIOS study comprises a population-based sample of 10,004 Asian men and women living in Singapore. Longitudinal data were collected in a random subset of 235 individuals.

### Plasma metabolome reflects self-reported dietary exposure

Plasma metabolomics profiling by mass spectrometry revealed 1,055 distinct metabolites, of which 887 (80%) are structurally identified. We found 156 metabolites positively correlated with 37 foods and beverages after adjusting for age, sex and ethnicity (*r*=0.15-0.68, FDR<0.05; Fig. 2). These metabolites reflect habitual dietary intakes within the multi-ethnic study population and are primarily from xenobiotics, lipid and amino acid pathways (Fig. 2).

**Fig. 2.**
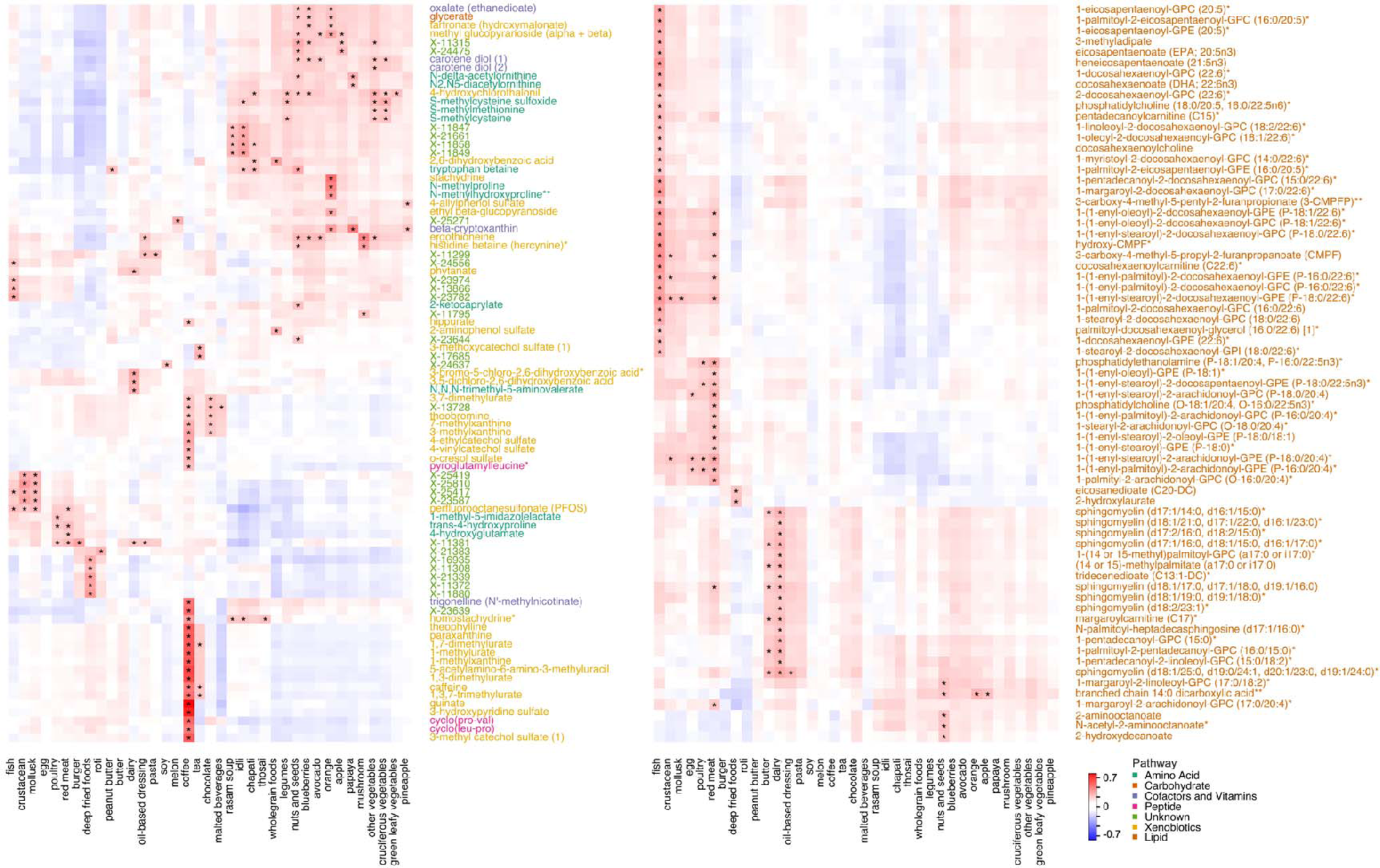
Plasma metabolome-wide association heatmaps with FFQ food and beverage items from partial correlation analysis, adjusted for age, sex and ethnicity. Left panel comprises of metabolites from amino acid, carbohydrate, cofactors and vitamins, peptide and xenobiotics pathways, and structurally unidentified metabolites; right panel comprises of metabolites from lipid pathways. *indicates FDR *P*<0.05, and correlation coefficients, *r*≥0.15. Ordering of variables were determined by hierarchical clustering.

We observed novel correlations for Asian ethnic-relevant foods, processed foods, fruits and vegetables, red meat, seafood, and nuts and seeds. Chapati, idli, thosai and rasam soup, which are frequently consumed by South Asians, were uniquely correlated with betainised compounds (homostachydrine and tryptophan betaine; *P*≤1.13×10^−7^) and other metabolites (X-11849, X-11858, X-21661, X-11847 and X-23639; *P*≤1.75×10^−4^). 4-Hydroxychlorothalonil, a metabolite of crop fungicide, Chlorothalonil, widely used to manage fungal diseases in agricultural crops^14^, was correlated with plant-based foods including vegetables, legumes, nuts and seeds (*P*≤2.05×10^−7^). Novel correlations were observed for roti (X-21383; *P*=2.52×10^−13^), apple (branched chain 14:0 dicarboxylic acid, methyl glucopyranoside, X-11315 and X-24475; *P*≤1.71×10^−7^) and tropical fruits including melon (X-25271; *P*=1.59×10^−14^), pineapple (4-allylphenol sulfate; *P*=2.84×10^−7^) and papaya (N2,N5-diacetylornithine and N-delta-acetylornithine; *P*≤3.94×10^−8^). N2,N5-diacetylornithine was previously reported elevated in blood after intakes of legumes^15^ and dry-bean enriched^16^ diets. X-11381 was correlated with burger and red meat (*r*=0.20-0.25; *P*≤6.0×10^−12^), similar to previous reports with red meat and milk^17^.

Deep fried foods correlated with fatty acids 2-hydroxylaurate and eicosanedioate (*P*≤3.15×10^−8^) and other metabolites (X-11880, X-11372, X-16935, X-21383 and X-21339; *P*≤1.06^−11^) previously associated with french fries, chips and fried food in 2 US and UK cohorts^17,18^. Specific lipid classes were characteristic for red meat, poultry and egg including phosphatidylethanolamines and plasmalogens containing stearoyl moiety (*P*≤4.72×10^−6^), which were distinct from the sphingomyelins, phosphatidylcholines, lysophospholipids and fatty acids characteristic for dairy products (*P*≤4.96×10^−7^, Fig. 2), of which tridecenedioate and sphingomyelin (d17:1/16:0, d18:1/15:0, d16:1/17:0) were previously associated with cheese^17^. Phytate, derived through ruminant metabolism of chlorophyl^19^, correlated with dairy products (*r*=0.18, *P*=3.0×10^−10^). Nuts and seeds correlated with 2-hydroxydecanoate, 1-margaroyl-2-linoleoyl-GPC and amino fatty acids (N-acetyl-2-aminooctanoate and N-acetyl-2-aminooctanoate; *P*≤1.09×10^−8^).

We also observed multiple known biomarkers of fish, coffee, cruciferous vegetables, mushroom and orange in our study, validating their relevance in our multi-ethnic Asian population. Fish had the most correlations (39 metabolites), where 3-carboxy-4-methyl-5-propyl-2-furanpropanoate (CMPF)^20,21^ and hydroxy-CMPF^22^ were most strongly correlated (*r*>0.30; *P*≤6.87×10^−28^), followed by various n3 and n6 PUFA^21^, long chain acylcarnitines, lysophospholipids and plasmalogens containing docosahexaenoic acid (DHA) or eicosapentaenoic acid (EPA) moiety (*P*≤5.23×10^−7^). Novel fish-related correlations included X-13866, X-25417, X-23974 and X-23782 (*P*≤2.50^−9^). Seafood is generally categorised as a broad food group but in this study, was categorised separately to reflect different organisms. Distinct correlations for crustacean and mollusk were observed for X-23587, X-25810 and X25419 (*P*≤2.56×10^−11^).

Coffee had the next most correlations (27 metabolites; *P*≤4.14×10^−7^) with caffeine, xanthines, alkaloids (trigonelline), phenolic acids and diketopiperazines (cyclo(leu-pro) and cyclo(pro-val)) previously reported in global cohort studies^9,13,23^ and coffee itself^23,24^. X-23639 was reported for the first time in this population (*P*=6.12×10^−51^). X-17685, X-13728 and X-11795 were uniquely correlated to tea (*P*=1.37×10^−13^), chocolate-containing products (*P*=1.68×10^−7^) and mushroom (*P*=2.42×10^−8^), respectively.

Other known biomarkers replicated in our study population included ergothioneine and its metabolite, hercynine (*P*≤1.13×10^−20^) for mushroom^25,26^; sulfur-containing amino acids (S-methylcysteine sulfoxide, S-methylcysteine and S-methylmethionine; *P*≤2.30×10^−7^) for cruciferous vegetables^17,27^; proline betaine, N-methylproline and N-methylhydroxyproline (*P*≤2.63×10^−24^) for orange/citrus fruits^28–31^; β-cryptoxanthin (*P*≤2.86×10^−8^) for orange, pineapple and papaya^32,33^; 2-aminophenol sulfate (microbial metabolite of benzoxazinoids found in wheat and rye^34^) for wholegrain foods (*P*=1.22×10^−14^); 3-bromo-5-chloro-2,6-dihydroxybenzoic acid, 3,5-dichloro-2,6-dihydroxybenzoic acid and N,N,N-trimethyl-5-aminovalerate (*P*≤2.91×10^−13^) for dairy/milk^17,21^.

### Dietary biomarker panels comprise shared and distinct metabolite networks

To demonstrate the potential for plasma metabolites to be used as objective assessment of dietary intakes in our Asian epidemiological setting, we developed candidate dietary multi-biomarker panels for key foods and beverages using elastic net penalisation as a machine-learning feature selection approach. These biomarker panels were curated for specific foods (e.g., orange), broader food groups (e.g., fruits) and complex mixed dishes (e.g., noodle dishes) based on the available FFQ questions and achieving model variance adjusted-*R*^2^>0.1 (to minimise less precise predictions) in respective training sets.

These panels each consisted of 3-39 metabolites (280 total metabolites, of which 211 were unique), primarily from lipid (30%), xenobiotics (16%) and amino acid (16%) pathways (Fig. 3a). The panels comprised a combination of i) known diet-derived metabolites (e.g., *CMPF for fish, proline betaine for orange*), ii) metabolites incorporated into endogenous metabolism (e.g., *1-(1-enyl-stearoyl)-2-arachidonoyl-GPE (P-18:0/20:4) for red meat, 1-palmityl-2-arachidonoyl-GPC (O-16:0/20:4) for processed meat*), as well as 51 structurally unidentified metabolites that were iii) previously reported (4%) or iv) potentially novel (22%, Fig. 3b). Notably, metabolite panels for mollusk, poultry and red meat were partially composed of per- and polyfluoroalkyl substances (PFAS) including perfluorooctanesulfonate (PFOS) and perfluorooctanoate (PFOA), persistent environmental contaminants recognised as ‘forever chemicals’^35^, while metabolite panels for cruciferous vegetables, legumes, mushroom and wholegrain foods comprised 4-hydroxychlorothalonil; consistent with positive partial correlation results.

**Fig. 3.**
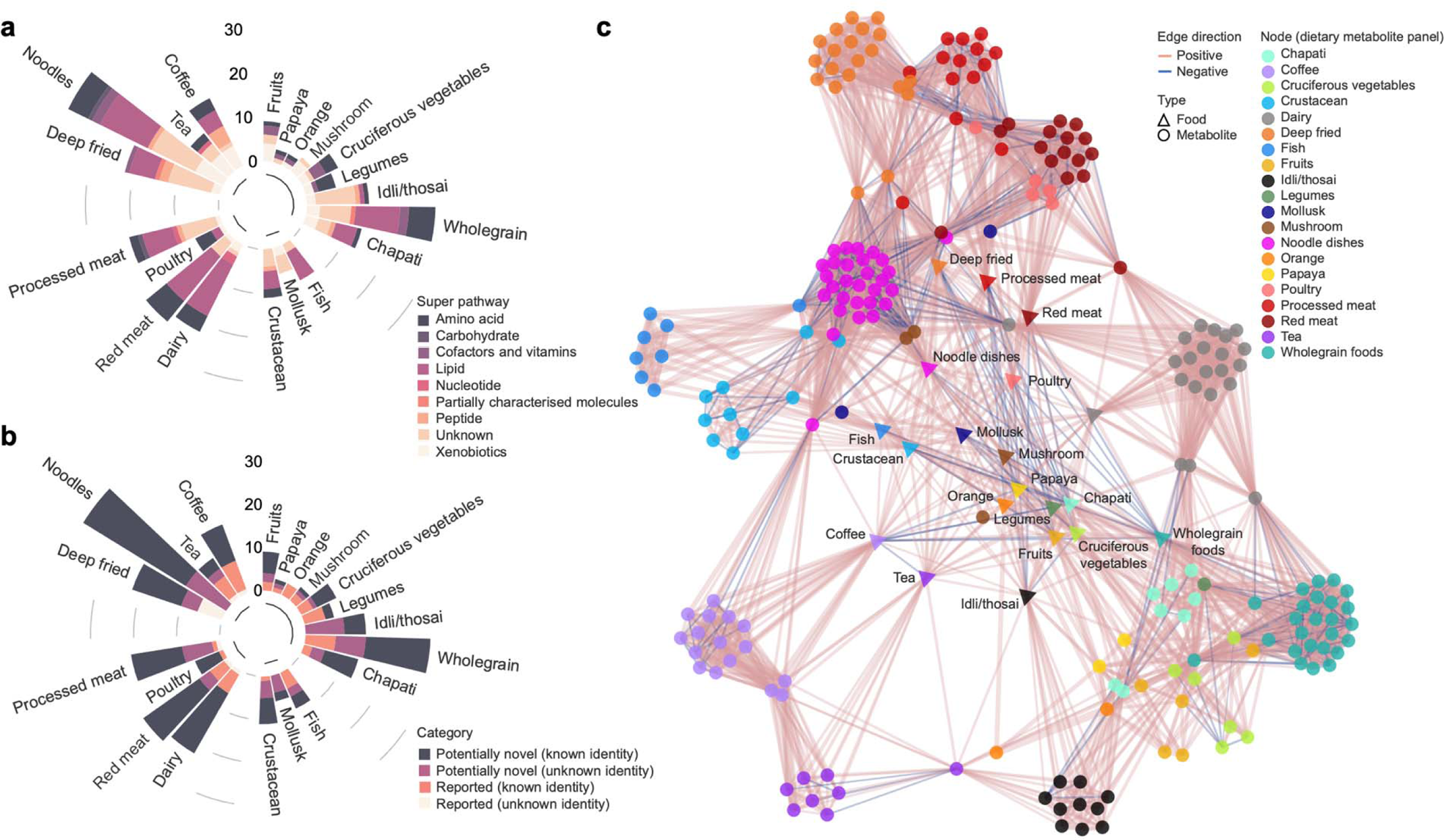
Candidate dietary multi-biomarker panels for foods and beverages a) assigned to super pathways, and b) categorised into reported or potentially novel biomarkers. C) Network graph summarising within and between dietary variables (foods and beverages) and metabolites derived from the biomarker panels reflecting respective dietary intakes, based on partial correlation coefficients (spearman *r*) adjusted for age, sex and ethnicity. Each node represents a food, beverage or metabolite. Line/edge colour represents direction of correlation (red for positive, blue for negative) and line/edge thickness represents strength of correlation.

Metabolic variation across different foods and beverages showed unique and shared networks (Fig. 3c). Red meat, processed meat, poultry and deep fried foods shared highly correlated networks, including 2R,3R-dihydroxybutyrate (formed via degradation of di/polysaccharides during cooking), X-11299 (associated with processed meat^17^), X-11880, X-16935, X21383 (associated with french fries and/or chips^17^) and 47 other metabolites. Ribitol, orotidine and a sulfated piperine metabolite (alkaloid in black/long pepper^36^) were distinct to processed meat panel. Margaroylcarnitine (C17), various plasmalogens, X-12822 and X-22776 were distinct to red meat panel, while gluconate, various peptides and lipids, X-11308, X-11372, X-21339 and X-23680 were unique to deep fried panel.

The meat-related foods were linked, via eicosanedioate (C20-DC), N-stearoyl-sphingosine (d18:1/18:0), X-23662 and X-07765, to noodle dishes as a mixed complex food group, typically composed of noodles with minor heterogenous proportions of seafood, animal and/or plant-based ingredients. This reflects its metabolite panel, being largest (39 metabolites) of the foods/beverages, distinctly including a glucuronidated piperine metabolite, S-allycysteine (biomarker of garlic^37^), androgenic steroids (reflective of animal organs intake), delta-CEHC (metabolite of tocopherol, a fortified ingredient in Asian wheat noodles^38^), anthranilate, X-13507, X-235937, X-11843, X-26106 and X-21310. Noodle dishes also shared networks with mushroom via ergothioneine and hercynine, with seafood via hydroxy-CMPF, X-23587, X-25417 and X-25810, and with coffee via γ-glutamylvaline.

Besides sharing 3 caffeine-related networks with coffee, the tea panel distinctly comprised N1-methyladenosine, N-lactoyl valine (formed from free amino acids and lactic acid in fermented foods^39^, and also associated with dairy (*P*=2.08×10^−5^)), thymol sulfate, 5-hydroxyindole sulfate, X-17685 and X-23654. Tea correlated highly with idli/thosai (*r*=0.68) and connected via X-18901. South Asian-ethnic foods shared 3 networks via X-13658, homostachydrine and 2,6-dihydroxybenzoic acid, of which the latter 2 were also present in wholegrain panel. Various dicarboxylate-, polyunsaturated-, long- and medium-chain fatty acids, amino acids, X-17654, X-24494, X-14939, X18799, X-21467 and X11478 were unique to wholegrain panel. The cruciferous vegetable panel shared common networks with legumes via S-methylcysteine sulfoxide, S-methylmethionine, carotene diol (2), 4-hydroxychlorothalonil, and with fruits via 4-allylphenol sulfate, a non-specific microbial metabolite of polyphenols^40^. We developed metabolite panels for fruits as a broad category of 17 fruits, and specifically for orange and papaya, which had strong discriminating metabolites (X-25271 for papaya, X-24475 for fruits).

Dairy panel shared 6 networks with red meat via X-11381, with noodle dishes via ceramide (d18:1/14:0, d16:1/16:0), with wholegrain via 10-undecenoate (11:1n1), with chapati via tridecenedioate (C13:1-DC) and heptenedioate (C7:1-DC), while having 16 distinct metabolites including phytanate, 3-bromo-5-chloro-2,6-dihydroxybenzoic acid, N,N,N-trimethyl-5-aminovalerate and 1-(14 or 15-methyl)palmitoyl-GPC (a17:0 or i17:0).

### Multi-biomarker panels improve prediction of dietary intakes

We evaluated the effectiveness of our multi-biomarker panels in estimating dietary intakes in respective test sets using regression models adjusted for age, sex, ethnicity (model 1), additionally for income and years of education (model 2), and multi-biomarker panels (model 3). Models with multi-biomarker panels accounted for 3-48% higher variances (*P*≤1.95×10^−19^; Fig. 4a). Coffee panel achieved the most variance (51%, *P*=2.23×10^−308^), followed by idli/thosai panel (50%, *P*=5.79×10^−42^) and chapati panel (43%, *P*=5.12×10^−37^). The latter two foods strongly reflected ethnic-specific foods, and in combination with legumes, are frequently consumed by South Asians, compared to Chinese and Malay populations. This finding is consistent with (South Asian) ethnicity being a major contributor to the variance for these foods (idli/thosai: β=1.70±0.69, *P*=2.12×10^−120^; chapati: β=0.95±0.05, *P*=3.17×10^−78^; legumes: β=0.64±0.05, *P*=6.96×10^−32^). These multi-biomarker panels explained 0.02-7% more variance of intakes (*P*≤2.23×10^−4^) compared to single biomarkers (with highest variance; Extended Fig. 1).

**Fig. 4.**
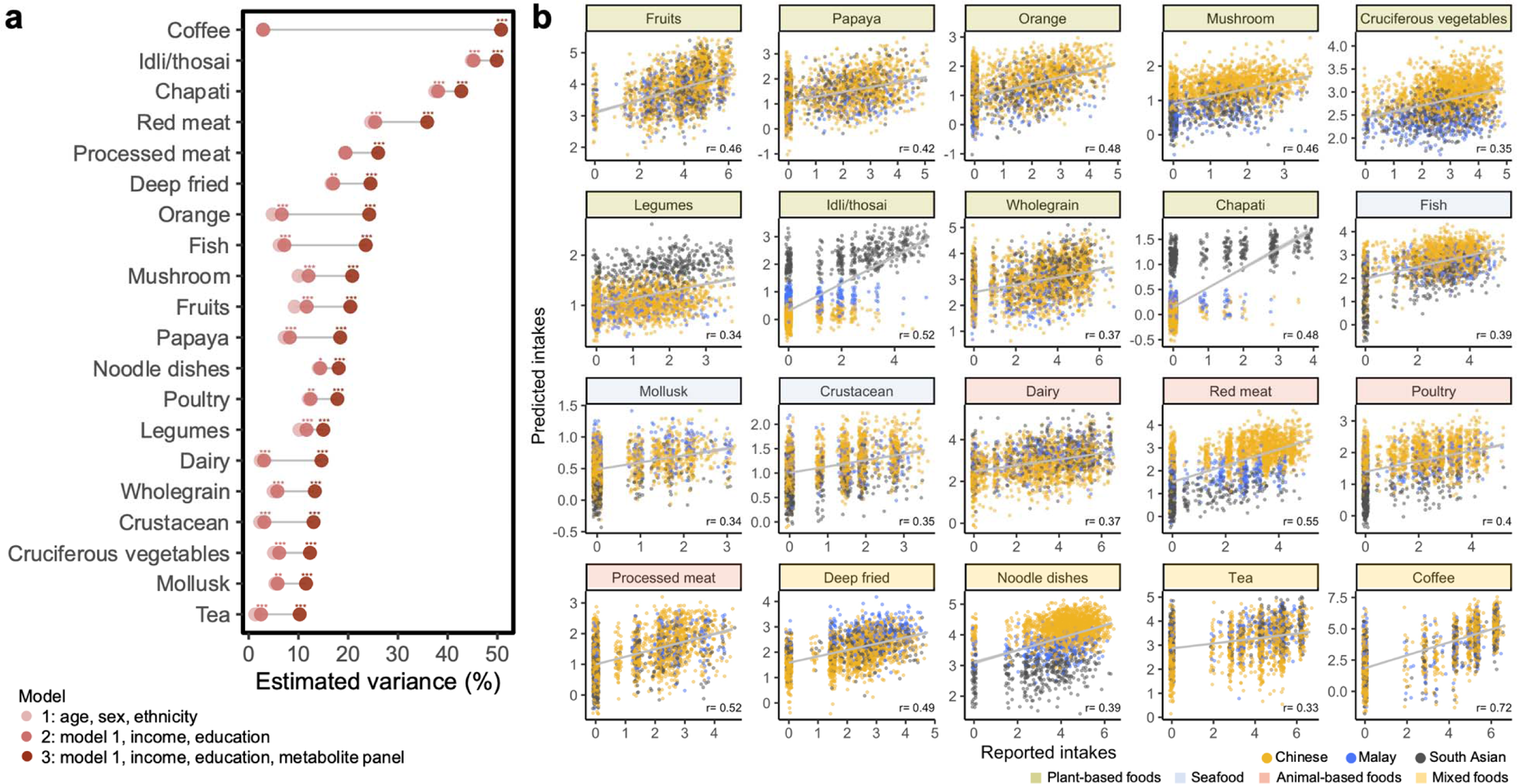
A) Dietary multi-biomarker panels improved prediction of intakes of foods and beverages in independent regression models of respective test sets (sample sizes for test sets in Extended Data Table 3). Values represent proportion of variance estimated. Significant difference between models (model 1 vs 2; model 2 vs 3) is represented by \*\**P*<0.01, \*\*\**P*<0.001. b) Scatter plots showing agreement (spearman *r*) between self-reported (x-axis) vs predicted (y-axis) intakes (log-transformed) for each food and beverage. Each point represents an individual and are coloured by their reported ethnicity.

Estimated dietary intakes, derived from these panels showed moderate to good agreement and ranking validity with self-reported FFQ intakes, *r*=0.33-0.72 (Fig. 4b). Error was greatest amongst individuals reporting zero intake and excluding this group of individuals improved median prediction accuracy (Extended Data Fig. 2). Coffee had highest prediction accuracy (84%), compared to other foods, but accuracy was also high (>70%) for meat, fish, fruits, cruciferous vegetables and noodle dishes. This demonstrates the utility of biomarker panels as a more accurate dietary assessment, compared to self-reporting tools.

### Utilising metabolite scores to investigate association with clinical phenotypes

We developed metabolite composite scores for each food and beverage, as a weighted sum of metabolites from the dietary biomarker panels. Compared to self-reported intakes, the scores associated significantly and more strongly with clinical phenotypes including homeostatic model assessment for insulin resistance (HOMA-IR), type 2 diabetes (T2D), BMI, fat mass index (FMI), carotid intima-media thickness (cIMT) and hypertension (Fig. 5). Scores of ‘healthy’ plant-based foods including fruits, vegetables, wholegrain foods and chapati as well as fish and dairy products were negatively associated with these phenotypes, with the third tertile corresponding to the strongest associations (*P*=2.39×10^−119^–2.37×10^−2^), as expected and consistent with the direction of associations based on self-reporting (*P*=3.03×10^−16^-5.91×10^−3^). For glycemic and adiposity indices, fruits scores showed the most protective effect (HOMA-IR: β=−0.60±0.03, *P*=4.34×10^−110^; T2D: β=−1.37±0.10, *P*=1.78×10^−45^; BMI: β=−0.56±0.03, *P*=7.21×10^−104^; FMI: β=−0.57±0.02, *P*=2.39×10^−119^). For vascular disease risks, legume scores showed the most protective effect (cIMT: β=−0.22±0.03, *P*=2.29×10^−18^; hypertension: β=−0.59±0.07, *P*=5.62×10^−17^).

**Fig. 5.**
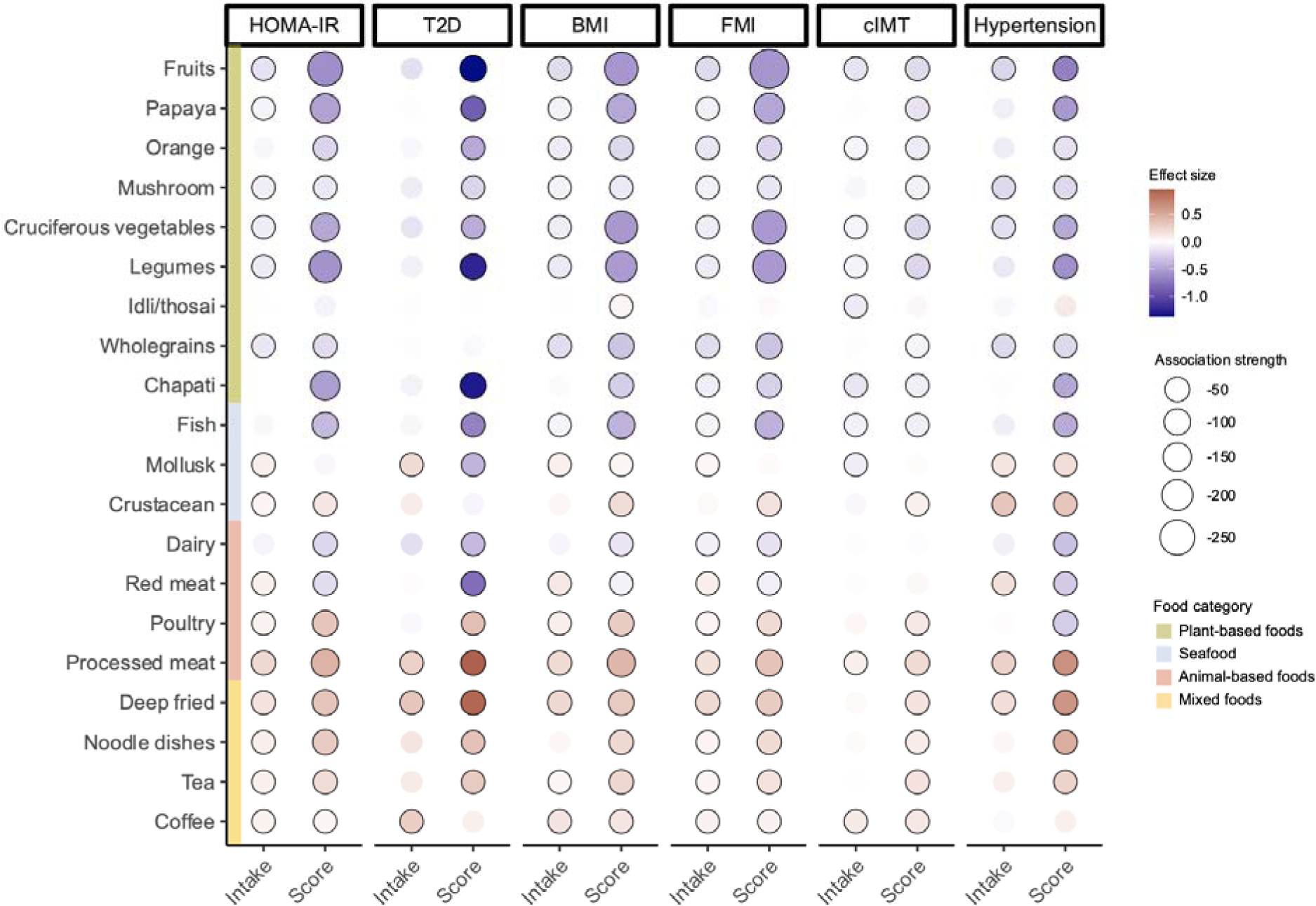
Associations between clinical phenotypes and third tertiles of dietary intakes derived from FFQ-based self-reported intake or metabolite score, adjusted for age, sex and ethnicity (n=8,391), with first tertile (either intake or score) as reference. β-coefficients were obtained from independent linear regression (for HOMA-IR, BMI, FMI, cIMT) or logistic regression (for T2D, hypertension) for each food/beverage, based on the following sample sizes: T2D (n=1,040 T2D, n=7,351 non-T2D), hypertension (n=2,411 with hypertension, n=5,980 without hypertension). Association strength represents log-transformed *P* values. Circle border represents significant associations (*P*<0.05). HOMA-IR, homeostatic model assessment for insulin resistance; T2D, type 2 diabetes; FMI, fat mass index; cIMT, carotid intima-media thickness.

In contrast, other seafood, animal-based and processed foods associated positively with these cardiometabolic phenotypes (scores: *P*=1.05×10^−61^-3.08×10^−2^; reported intakes: *P*=6.67×10^−19^-3.61×10^−2^). Amongst the ‘unhealthy’ foods, processed meat scores showed the most detrimental effect (HOMA-IR: β=0.45±0.03, *P*=6.53×10^−60^; T2D: β=0.95±0.09, *P*=6.27×10^−24^; BMI: β=0.43±0.03, *P*=1.05×10^−61^; FMI: β=0.35±0.02, *P*=1.69×10^−45^; cIMT: β=0.22±0.03, *P*=7.22×10^−18^; hypertension: β=0.65±0.03, *P*=2.38×10^−21^). Red meat score associated negatively with most phenotypes except cIMT (*P*≤1.15×10^−2^), contrasting the positive associations from self-reported intakes in our study (*P*≤1.25×10^−2^) and meta-analysis of 3 US cohorts^41^, suggesting preparation methods^42^ and biomarker composition play a role in influencing health associations. The red meat panel comprised primarily PUFA, and lesser proportions of amino acids, and *trans* and saturated fatty acids (‘unhealthy fats’), likely contributing to overall negative associations.

Coffee and tea intakes, derived from both scores and self-reported intakes, were positively associated with most phenotypes (*P*≤3.10×10^−2^), considering the trend of intakes in this study population being predominately energy-dense variations (with condensed/evaporated/whole milk, sugar or 3-in-1, Extended Data Fig. 3). Overall, the associations with metabolite scores were robust to BMI as a mediator, unlike the associations with self-reported intakes, which were no longer significant (Extended Data Fig. 4).

### Diet-metabolite relationships are robust across multiple visits

We evaluated the temporal consistency of diet-metabolite relationships in the subset of individuals who returned for a repeat visit. Plasma metabolites of the dietary biomarker panels from the first visit were present also in plasma profiles at the repeat visit, showing stability of these metabolites and diet-associations over time. Using prediction models of biomarker panels trained in the first visit, we estimated intakes of respective foods and beverages at the repeat visit, which showed good agreement with self-reported intakes recorded at the repeat visit, *r*=0.35-0.73 (Fig. 6a). Metabolite scores also showed good agreement between first and repeat visits, *r*=0.46-0.80 (Extended Data Fig. 5a).

**Fig. 6.**
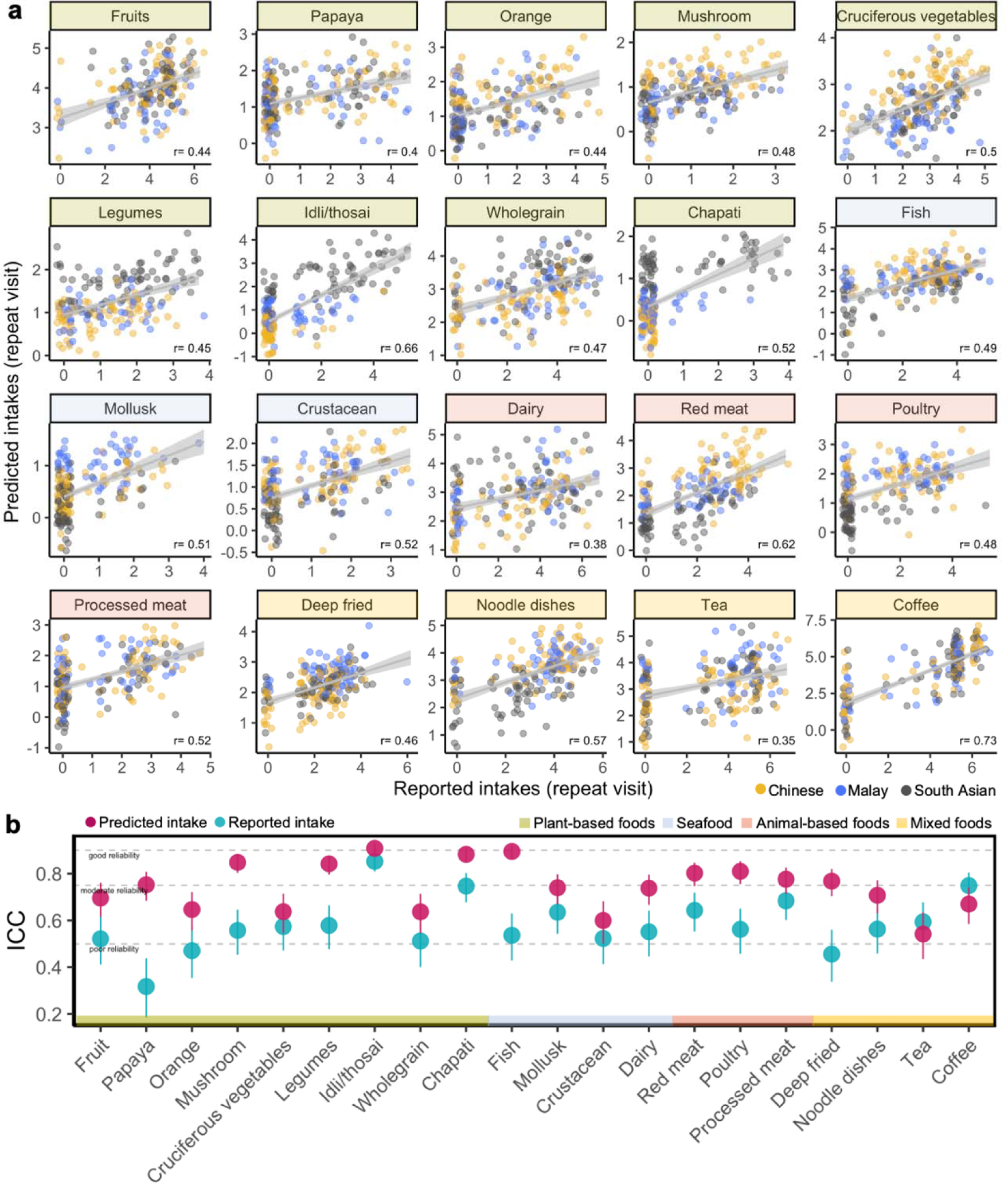
A) Scatter plots showing agreement (spearman *r*) between self-reported (x-axis) vs predicted (y-axis) intakes (log-transformed) for each food and beverage in the repeat visit (n=195 pairs of individuals); predicted intakes were derived from models trained in training sets from first visit sets. Each point represents an individual and are coloured by their reported ethnicity. B) Intraclass correlation coefficients (ICC) showing reliability of reported and predicted intakes across first and repeat visits.

Across first and repeat visits, the reliability and reproducibility of predicted intakes (intraclass correlation coefficients (ICC)=0.54-0.91) consistently outperformed the self-reported intakes (ICC=0.32-0.85), with 50% of foods and beverages achieving ICC>0.75, indicating good reliability (Fig. 6b). Similarly, metabolite scores showed good reliability (ICC=0.50-0.89; Extended Data Fig. 5b).

## Discussion

We generated fasting plasma profiles of 1,055 metabolites in 8,391 individuals (235 with repeat data) for whom we had extensive dietary records, sociodemographic, health and lifestyle variables. We carried out diet-metabolite association analyses and developed 20 dietary biomarker panels that were representative of food and beverage intakes within a multi-ethnic Asian population, using machine-learning based prediction models for each intake. We showed the feasibility and application of these biomarker panels to objectively assess dietary intakes, compared to self-reports, whose limitations are well-recorded.

Biomarker-calibrated models for single foods^12,43,44^ based on highly specific biomarkers and broader metabolomics profiles characterising dietary patterns (DASH^44^ and HEI^45^) have been reported in American and European populations but not developed or validated in other populations. Considering the chemical complexity of diets in different regions, our study highlights the importance of mapping the highly individualised Asian metabolome in a geographically and culturally relevant context. When used to predict daily intakes, some of these biomarker panels captured ethnic differences in dietary habits (e.g., idli/thosai and noodle dishes), which is key to documenting diets underlining a multi-ethnic population. Metabolite-based modelling represents an avenue to explore, particularly for culturally-relevant foods that are under-studied, compared to common/Westernised foods.

Metabolites of these dietary biomarker panels were derived from dietary or environment sources as well as compounds incorporated into metabolic or physiological processes. We showed that using a combination of metabolites (as a panel) explained higher variance of dietary intakes and significantly improved intake predictions, compared to models composed only of sociodemographic variables. Dietary intakes estimated by an individual’s plasma biomarker panel correlated with self-reports for specific foods or complex dishes, and these food-metabolite relationships were stable over time. Apart from the utility as an intake prediction tool, the panels also behaved as a proxy of intakes that improved accuracy of clinical associations.

We acknowledge our study is observational with most individuals having dietary and metabolite datasets at one timepoint and the metabolite-phenotype associations do not highlight causal relationships, a similar limitation encountered by other epidemiological studies. Plasma metabolite levels mirror recent diet history and may not reflect long-term kinetics beyond the metabolites half-times (few hours to days) depending on frequency of intakes. Some metabolites within the panels are structurally unidentified and hence not well-studied in the context of their derived source or chemistry, and should be interpretated with caution. Our novel metabolite associations and panels, particularly for ethnic-specific foods will benefit from replication in other Asian populations.

Our multi-ethnic Asian population study provides new insights into diet-related metabolic variations, including ethnic-specific dietary habits and environmental effects on the food supply. We demonstrate that the application of dietary biomarker panels, objectively representative of food and beverage intakes in complex diets, is feasible, and generates clinically-relevant insights. Our research provides a pathway to new opportunities to assess interindividual variability in metabolism and discriminate metabotypes, and to link exposure to health outcomes in Asian populations.

## Methods

### General cohort information

Datasets were derived from the Health for Life in Singapore (HELIOS) study (ethics approval IRB-2016-11-030, www.healthforlife.sg), a multi-ethnic prospective cohort of adults aged 30-84 years old^46^. In HELIOS, 10,004 individuals (Singapore citizens or permanent residents) were recruited from the general population across socio-economic backgrounds and excluded if they were pregnant, breastfeeding or had prior major illness. Consenting individuals underwent extensive questionnaire, physiological, imaging and biological sample phenotyping. Individuals’ age, sex and ethnicity were recorded as per their identity card. Environmental and lifestyle information including income level, years of education, cigarette smoking and medical history were recorded using self-administered questionnaires. In this study, we excluded individuals whose ethnicity were not of the primary ethnic groups (Chinese, Malay, South Asian) or who did not complete physiological or biochemical assessments, FFQ or reported implausible total energy intake, or who did not have metabolomics data (Fig. 1).

### Clinical phenotyping

Fasted blood biochemistry including fasting glucose, HbA1c, total cholesterol, LDL, HDL and triglycerides were measured by local accredited laboratory (QuestLab Singapore, SAC-SINGLAS ISO 15189:2012). Insulin was measured in-house using ADVIA Centaur XP Immunoassay System (Siemens Healthcare, Erlangen, Germany). HOMA-IR is calculated as insulin (mIU/L)*glucose (mM)/22.1512. FMI was derived from body fat mass (kg) quantified using whole-body Dual X-ray Absorptiometry (DEXA) scans (Horizon^TM^ W densitometer S/N 30052M, Hologic, Massachusetts, USA, QDR software version 13.6.0.5) and normalised against height (m^2^). cIMT, a subclinical atherosclerotic phenotype, was estimated from 2D and 3D carotid ultrasound imaging of the common carotid artery using Philips iU 22 ultrasound system. cIMT measurement was centered at 1 cm proximal to the carotid bifurcation; images were obtained at the lateral and posterior angles bilaterally, and analysed using Philips Qlab software. Measurements were quality checked to ensure ≥95% of tracing were correctly identified and coefficient of variation was < 5% for inter-operator and inter-reader variability. Mean cIMT values were natural log-transformed for downstream analyses. Individuals were classified with T2D based on ≥7 mM fasting glucose level, ≥6.5% HbA1C or prior diagnosis by doctor (self-reported history). Individuals were classified with hypertension based on systolic blood pressure (SBP) ≥140 mmHg, diastolic blood pressure (DBP) ≥90 mmHg, prior diagnosis by doctor (self-reported history) or taking hypertensive medications.

## Data generation and preprocessing

### Dietary assessment

Dietary intakes were assessed by self-administered computerised Food Frequency Questionnaire (FFQ), which was validated in a local population^47^. The FFQ comprises of 169 local multi-ethnic food and beverage items with corresponding energy and nutrient composition database (Singapore Health Promotion Board, https://focos.hpb.gov.sg/eservices/ENCF). Total daily energy intakes were calculated for each individual and excluded if their energy intakes were considered implausible based recommended energy intake thresholds, i.e., <500 kcal/day or >3 SD of gender specific mean)^48^. Individual food or beverage items were grouped into broader food and beverage groups based on their biological classification and/or nutritional property (Extended Data Table 2). For each food/beverage group, intakes (grams/day) >99.5 percentile were considered outliers and excluded.

### Plasma metabolome

Untargeted metabolomics profiling of plasma samples was performed by Metabolon, Inc (North Carolina, USA)^49–51^ on fasting plasma samples. Briefly, samples were methanol-extracted and analysed by untargeted liquid chromatography-tandem mass spectrometry (LC-MS/MS) Metabolon Global Discovery platform using 2 reversed-phase (RP) methods in positive electrospray ionisation (ESI) mode, a RP method in negative ESI and a HILIC method in negative ESI, from 70-1000 m/z range. A cocktail of quality control (QC) standards was added to each sample (before injection) for monitoring of instrument performance and chromatographic alignment. Aliquots from each sample were pooled to generate a pooled matrix sample that was periodically injected as technical replicates and to distinguish between biological and process variability. Samples were randomised within batch runs. Raw semi-quantitative data of peaks was extracted and quantified from area-under-the-curve, identified, QC-processed and normalised across shipment batches using median-scaling. Annotated/identified metabolites were assigned to nine pathways corresponding to their broad chemical class. Metabolites of unknown structural identity were assigned numeric reference numbers beginning with X.

Five samples that failed QC standards were excluded. Metabolites with >20% missingness (assuming not detected) were excluded and the remaining were imputed with minimum value for each metabolite (assuming missingness due to low abundance). Metabolites were further excluded if they were considered outliers (defined as >6 SD away from the mean of the first 2 principal components in overall and platform-specific Principal Component Analysis. Median relative standard deviation (SD) of instrument and technical variability between batch runs ranged from 6-9% and 10-12%, respectively. Metabolite levels were natural log-transformed and scaled for downstream analyses.

## Statistical analysis

### Metabolome-wide correlation with dietary habits

To assess associations between diet variables and metabolites, partial correlation analysis was performed and controlled for effects of age, sex, ethnicity and batch. *P* values were FDR-adjusted with Benjamin-Hochberg method. To select meaningful associations, only spearman rank correlation coefficients, *r*≥0.15 that were significant (FDR<0.05) were considered.

### Estimating variance of individual metabolites

To estimate the variance of each metabolite that contributed to individual diet variables, we applied univariate OLS regression (*glmnet* package, version 4.1-7), adjusted for age, sex, ethnicity and batch. In all regression analyses, all numeric variables were natural log-transformed and scaled in advance. In all cases of multiple testing, *P* values were FDR-adjusted with Benjamin-Hochberg method and considered significant if *P*<0.05.

### Metabolite panels for predicting dietary intakes

Each food/beverage dataset was split into training (70%) and test (30%) sets based on similar proportions of respective food/beverage, age, sex and ethnicity (refer to Extended Data Table 3 for training and test sets’ sample sizes). As a variable selection method and for handling highly correlated predictors, we applied machine learning-based elastic net^52^ regression with 10-fold cross-validation (*glmnet* package, version 4.1-7) to each training set. Covariates (age, sex, ethnicity, income and years of education) were included in elastic net regression models without penalty factor (implying no shrinkage), unlike the metabolites whose coefficients were subjected to lambda, λ, penalty term. Optimal alpha, α, values were tuned at 2 levels, from 0 to 1 (resolution=0.1) and subsequently from ±0.05 of α (at the minimum mean-squared error, resolution=0.01). To achieve a parsimonious multi-biomarker panel for each food/beverage (outcome variable), metabolites derived from each model with non-zero positive coefficients were inspected and excluded from respective panels based on condition that they were not more strongly associated with other types of food/beverage. Partial correlations within and between metabolites (of the metabolite panels) and foods/beverages were summarised into graphical networks using fruchterman-reingold layout (*ggraph* package, version 2.1.0). The metabolite panels were evaluated for their effectiveness as a tool for predicting respective food and beverage intakes in the test sets based on their variance (adjusted *R*^2^>0.1) and spearman rank correlation coefficient, *r*, for agreement between reported and predicted intakes. Dietary variables were assigned to four categories corresponding to source and complexity (plant-based, animal-based, seafood and mixed foods).

### Metabolite scores for assessing phenotypes’ associations

Composite scores were developed for each multi-biomarker panel as an objective assessment of each food and beverage intake for each individual, by applying penalised weights derived from ridge regression coefficients to the metabolites abundance and taking the sum. The composite scores were subsequently categorised into tertiles and tested for associations with clinical phenotypes (HOMA-IR, T2D, BMI, FMI, cIMT and hypertension) in comparison to self-reported intakes (also categorised into tertiles) in independent OLS regression models (for HOMA-IR, BMI, FMI, cIMT) or logistic regression models (for T2D and hypertension), adjusted for age, sex and ethnicity. As BMI may be a potential confounder for the tested phenotypes, we performed sensitivity analysis including BMI as an additional covariate. Clinical data was scaled to allow comparison across phenotypes.

### Temporal consistency of individual metabolite/diet variables

There were 235 individuals for whom data and samples had been collected from 2 visits, 336 days apart on average, which were used to evaluate the temporal consistency of diet-metabolite relationships. After excluding 40 individuals with implausible daily energy intakes^48^, 195 individuals remained with data collected, 322 days apart on average (∼11 months). Prediction models, for each food and beverage, trained using data from the first visit was used to predict respective intakes in the repeat visit, and assessed for agreement between reported and predicted intakes via spearman rank correlation coefficient, *r*. The test-retest reliability within reported intakes, predicted intakes and metabolite scores across visits were determined via ICC using a two-way mixed effects model (type= consistency; *irr* package, version 0.84.1).

## Acknowledgements

We express our thanks to participants of the HELIOS study and the HELIOS operation team for recruitment, organisation and data/sample collection. This study (NTU IRB: 2016-11-030) is supported by Singapore Ministry of Health’s (MOH) National Medical Research Council (NMRC) under its OF-LCG funding scheme (MOH-000271-00), Singapore Translational Research (StaR) funding scheme (NMRC/StaR/0028/2017), the National Research Foundation, Singapore through the Singapore MOH NMRC and the Precision Health Research, Singapore (PRECISE) under the National Precision Medicine programme and intramural funding from Nanyang Technological University, Lee Kong Chian School of Medicine and the National Healthcare Group. Dorrain Low is also supported by the National Research Foundation, Singapore, through the Singapore MOH NMRC and PRECISE under the National Precision Medicine programme. Parts of figures were created with Biorender.com.

## Author contributions

J.C. J.B., P.E., E.R., J.L., E.S.L. and J.N. acquired funding for HELIOS study. J.C., K.W., P.A.S., R.S. and G.M. acquired funding for metabolomics analysis. J.C. and D.Y.L. conceptualised the study. D.Y.L., T.H.M., N.S., P.R.J., R.D., K.W., P.A.S., R.S. and G.M. generated the data. D.Y.L., T.H.M., N.S. and P.R.J. analysed the data. J.C. and D.Y.L. wrote initial manuscript draft. M.L., T.H.M., N.S., P.R.J. and P.A.S. provided critical feedback to manuscript draft. All authors reviewed and revised the final manuscript.

## Data availability

Request to access the data is available via for consideration of HELIOS study principal investigators.

## Extended Data

### Extended Data Tables

**Extended Data Table 1.**
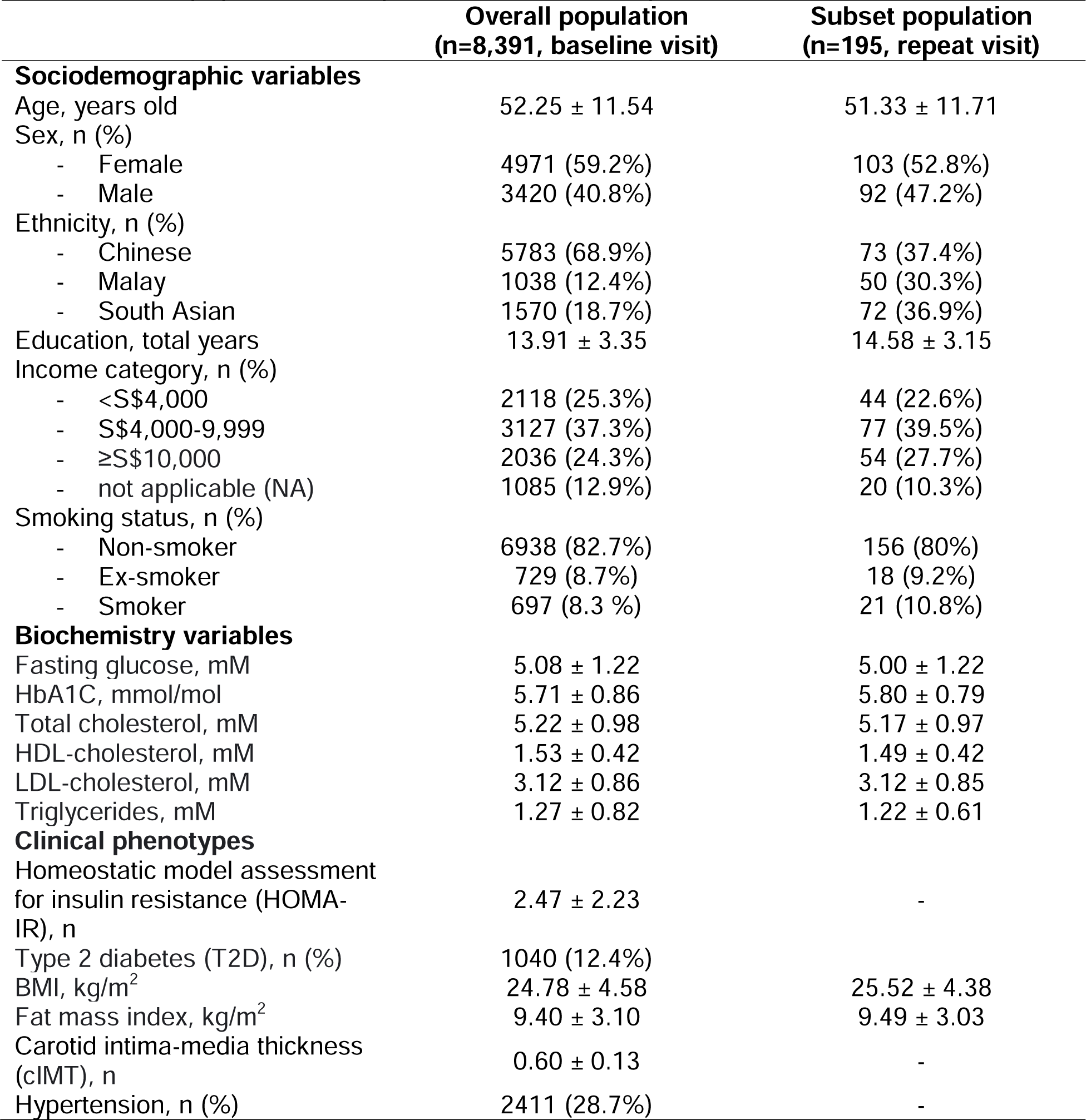
Characteristics of HELIOS study population at baseline visit and subset of population at repeat visit. Values are presented as mean ± SD.

|  | Overall population<br>(n=8,391, baseline visit) | Subset population<br>(n=195, repeat visit) |
| --- | --- | --- |
| **Sociodemographic variables** |  |  |
| Age, years old | 52.25 ± 11.54 | 51.33 ± 11.71 |
| Sex, n (%) |  |  |
| - Female | 4971 (59.2%) | 103 (52.8%) |
| - Male | 3420 (40.8%) | 92 (47.2%) |
| Ethnicity, n (%) |  |  |
| - Chinese | 5783 (68.9%) | 73 (37.4%) |
| - Malay | 1038 (12.4%) | 50 (30.3%) |
| - South Asian | 1570 (18.7%) | 72 (36.9%) |
| Education, total years | 13.91 ± 3.35 | 14.58 ± 3.15 |
| Income category, n (%) |  |  |
| - <S\$4,000 | 2118 (25.3%) | 44 (22.6%) |
| - S\$4,000-9,999 | 3127 (37.3%) | 77 (39.5%) |
| - ≥S\$10,000 | 2036 (24.3%) | 54 (27.7%) |
| - not applicable (NA) | 1085 (12.9%) | 20 (10.3%) |
| Smoking status, n (%) |  |  |
| - Non-smoker | 6938 (82.7%) | 156 (80%) |
| - Ex-smoker | 729 (8.7%) | 18 (9.2%) |
| - Smoker | 697 (8.3 %) | 21 (10.8%) |
| **Biochemistry variables** |  |  |
| Fasting glucose, mM | 5.08 ± 1.22 | 5.00 ± 1.22 |
| HbA1C, mmol/mol | 5.71 ± 0.86 | 5.80 ± 0.79 |
| Total cholesterol, mM | 5.22 ± 0.98 | 5.17 ± 0.97 |
| HDL-cholesterol, mM | 1.53 ± 0.42 | 1.49 ± 0.42 |
| LDL-cholesterol, mM | 3.12 ± 0.86 | 3.12 ± 0.85 |
| Triglycerides, mM | 1.27 ± 0.82 | 1.22 ± 0.61 |
| **Clinical phenotypes** |  |  |
| Homeostatic model assessment<br>for insulin resistance (HOMA-IR), n | 2.47 ± 2.23 | - |
| Type 2 diabetes (T2D), n (%) | 1040 (12.4%) |  |
| BMI, kg/m<sup>2</sup> | 24.78 ± 4.58 | 25.52 ± 4.38 |
| Fat mass index, kg/m<sup>2</sup> | 9.40 ± 3.10 | 9.49 ± 3.03 |
| Carotid intima-media thickness<br>(cIMT), n | 0.60 ± 0.13 | - |
| Hypertension, n (%) | 2411 (28.7%) | - |

**Extended Data Table 2.** Description of food and beverage groups

| **Foods and/or beverages** | **eFFQ items** |
| --- | --- |
| **Seafood:** |  |
| - Fish | raw fish, nigiri, dried fish, canned fish, white fish, oily fish |
| - Crustacean | prawn, crab |
| - Mollusk | squid, oyster |
| **Animal-based:** |  |
| - Poultry | poultry |
| - Red meat | fresh pork (fat, lean), fresh beef (fat, lean), mutton (fresh, lean) |
| - Processed meat | poultry ham, processed poultry, cured pork |
| - Dairy | butter, milk (non-fat, low-fat, whole), yoghurt (reduced fat, regular), cheese (reduced fat, regular) |
| **Plant-based:** |  |
| - Cruciferous vegetables | broccoli, cauliflower, chye sim, kailan, white cabbage, coleslaw |
| - Legumes | peas, long beans, other beans, dhal |
| - Mushroom | mushroom |
| - Fruits | orange, apple, avocado, banana, blueberries, dragonfruit, durian, grape, guava, kiwi, mango, melon, papaya, pear, pineapple, strawberries, dried fruit |
| - Orange | orange |
| - Papaya | papaya |
| - Wholegrain | oats, wholemeal bread, wholemeal pasta |
| - Chapati | chapati |
| - Idli and thosai | idli, thosai |
| **Mixed foods:** |  |
| - Noodle dishes | noodles in gravy, noodles in soup, dry noodles, fried noodles |
| - Deep fried | french fries, fried chips, deep fried sweet snacks, puri |
| - Coffee | coffee with the following: condensed milk, evaporated milk, whole milk, low-fat milk, skimmed milk, soya milk, creamer no sugar, 3in1, 2in1, no milk, bottled |
| - Tea | tea with the following: condensed milk, evaporated milk, whole milk, low-fat milk, skimmed milk, soya milk, creamer no sugar, 3in1, 2in1, no milk, bottled |

**Extended Data Table 3.** Summary of dietary multi-biomarker panels for the tested foods and beverages in respective training and test sets. For each food/beverage, intakes > 99.5 percentile were considered outliers and excluded before splitting into training (70%) and test (30%) sets based on similar proportions of age, sex, ethnicity and respective food/beverage.

| **Foods and/or beverages** | **No. of metabolites in panel** | **No. of potentially novel metabolites (known identity)** | **Training set (n)** | **Test set (n)** |
| --- | --- | --- | --- | --- |
| **Seafood:** |  |  |  |  |
| - Fish | 9 | 5 (2) | 5,809 | 2,487 |
| - Crustacean | 11 | 10 (6) | 5,793 | 2,490 |
| - Mollusk | 6 | 6 (2) | 5,790 | 2,490 |
| **Animal-based:** |  |  |  |  |
| - Poultry | 7 | 6 (6) | 5,788 | 2,488 |
| - Red meat | 24 | 23 (16) | 5,778 | 2,496 |
| - Processed meat | 21 | 19 (12) | 5,797 | 2,486 |
| - Dairy | 22 | 16 (16) | 5,803 | 2,492 |
| **Plant-based:** |  |  |  |  |
| - Cruciferous vegetables | 9 | 6 (5) | 5,801 | 2,495 |
| - Legumes | 7 | 2 (2) | 5,801 | 2,495 |
| - Mushroom | 4 | 2 (1) | 5,799 | 2,499 |
| - Fruits | 9 | 7 (5) | 5,798 | 2,498 |
| - Orange | 3 | 0 | 5,803 | 2,493 |
| - Papaya | 3 | 2 (1) | 5,804 | 2,492 |
| - Wholegrain | 29 | 22 (15) | 5,805 | 2,491 |
| - Chapati | 13 | 11 (8) | 5,803 | 2,494 |
| - Idli and thosai | 14 | 14 (5) | 5,800 | 2,496 |
| **Mixed foods:** |  |  |  |  |
| - Noodle dishes | 39 | 39 (28) | 5,799 | 2,496 |
| - Deep fried | 22 | 16 (12) | 5,804 | 2,492 |
| - Coffee | 17 | 9 (9) | 5,733 | 2,478 |
| - Tea | 11 | 6 (5) | 5,766 | 2,482 |

### Extended Data Figures

**Extended Data Fig. 1.**
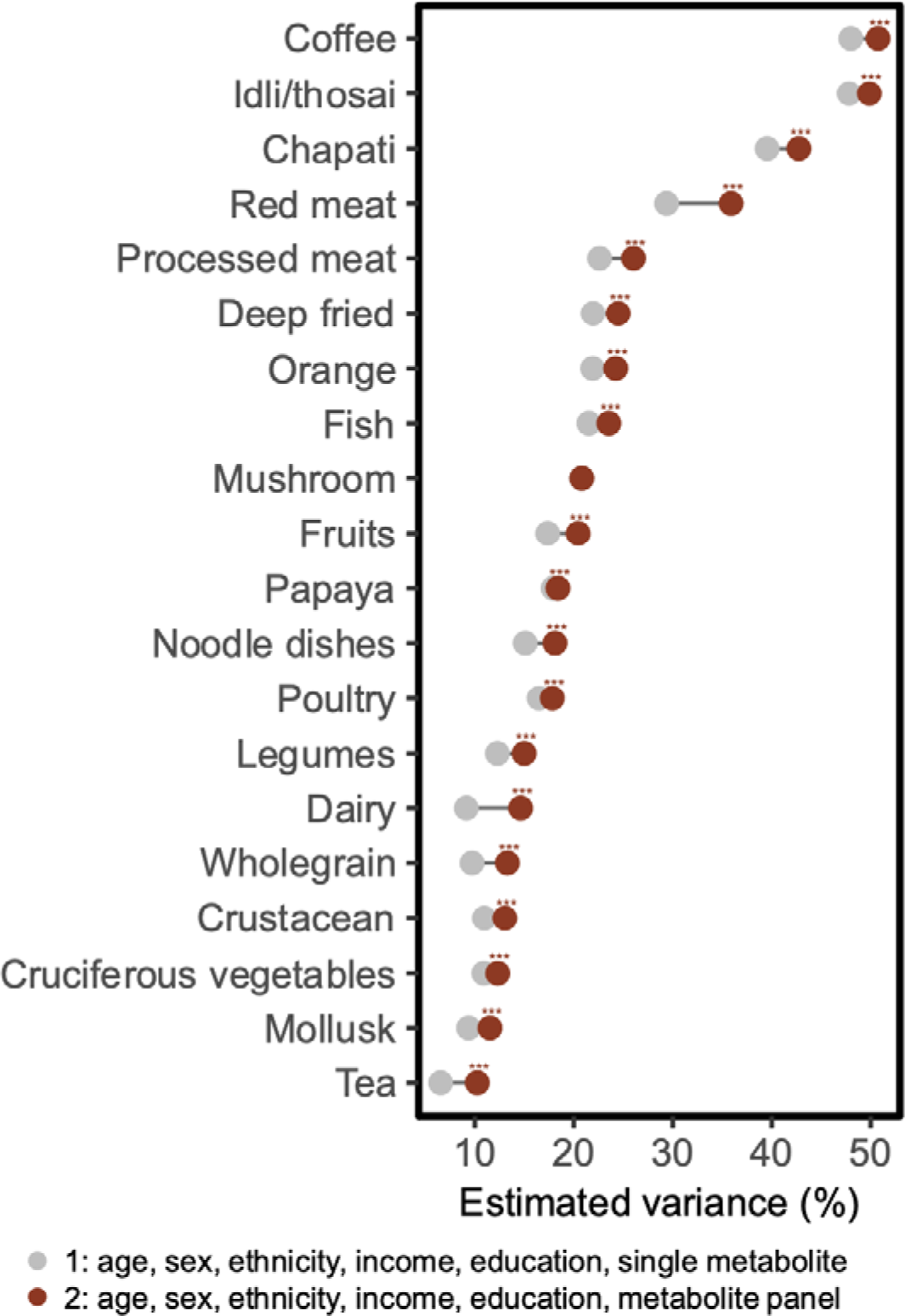
Dietary multi-biomarker panels significantly contribute more variance, compared to single metabolites, for respective food and beverage intakes. Significance difference between models is represented by \*\*\**P*<0.001 (*P* from 1.06×10^−12^-1.10×10^−111^).

**Extended Data Fig. 2.**
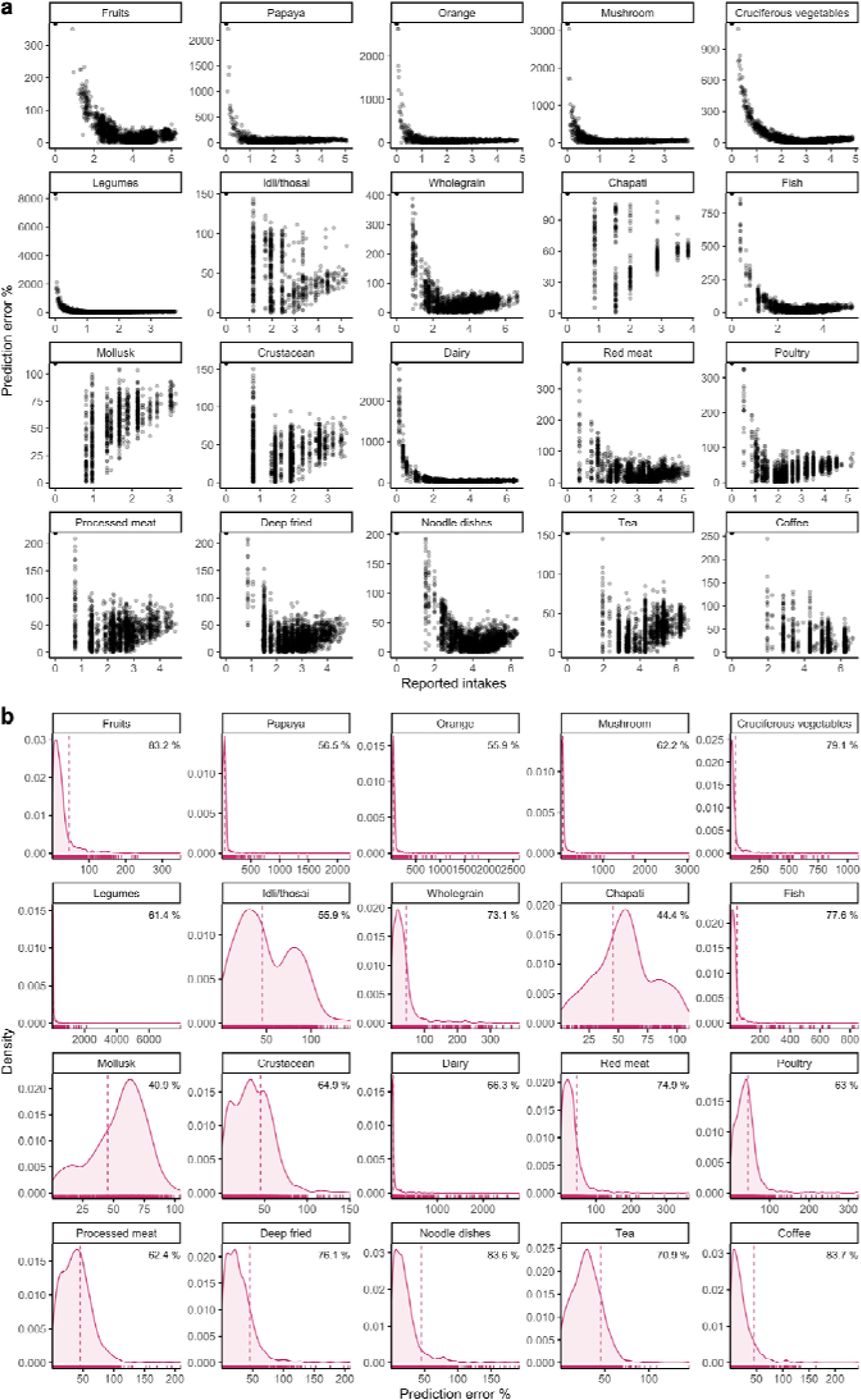
a) Percent (%) error observed between predicted and self-reported intakes (log-transformed) of foods and beverages in respective test sets. b) Density distribution of prediction error % of foods and beverages in respective test sets with dotted line indicating median error and text insert indicating median prediction accuracy (formula: 1 - error %).

**Extended Data Fig. 3.**
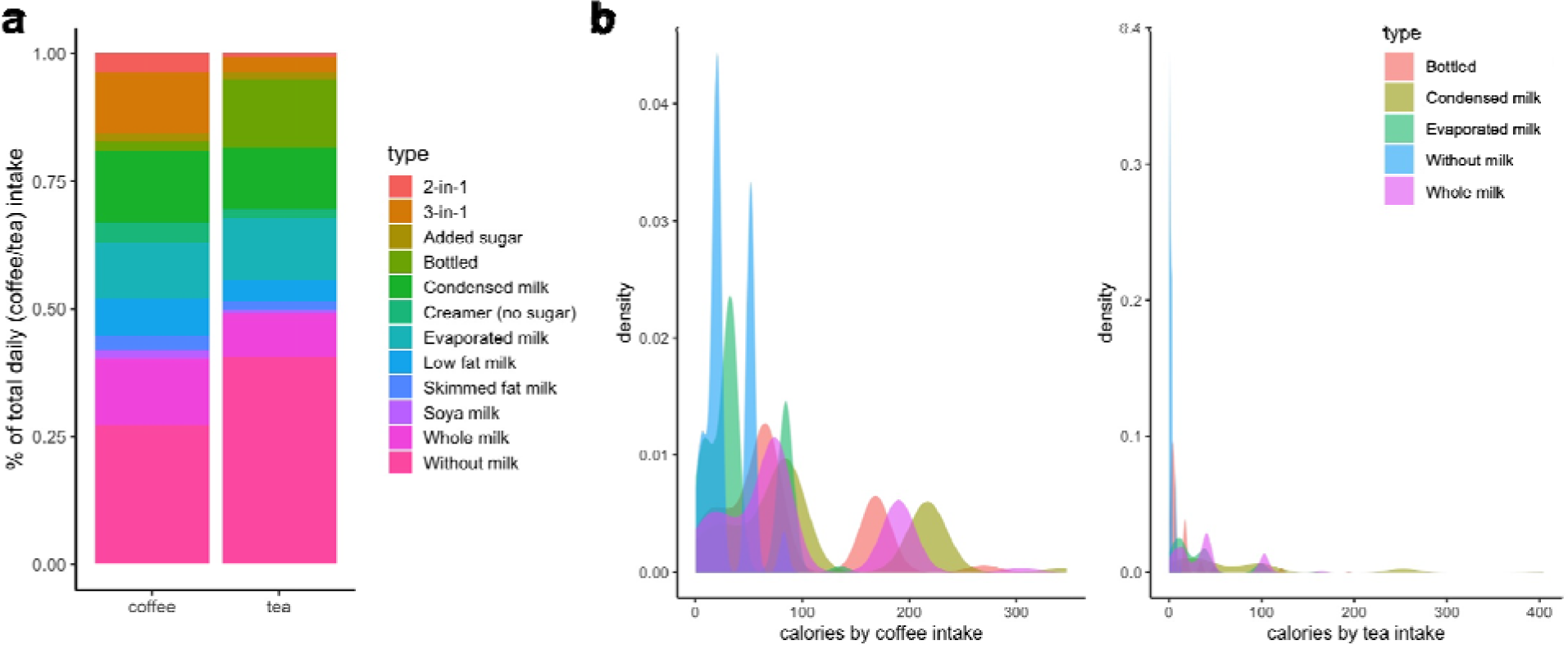
a) Percent (%) of variations of total daily coffee and tea intakes recorded via FFQ in the study population. b) Proportion of kilocalories across the top 5 most consumed variations of coffee or tea.

**Extended Data Fig. 4.**
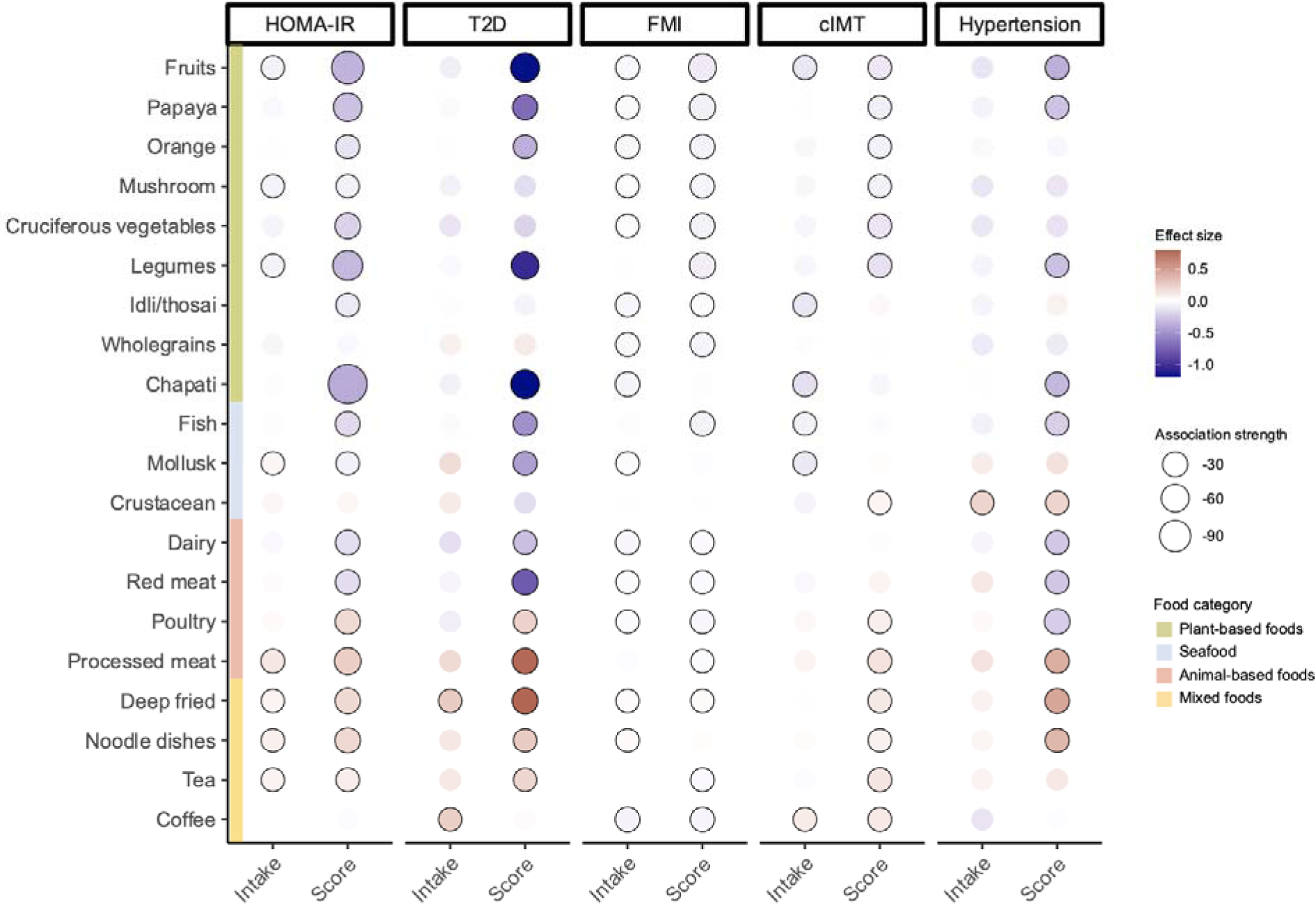
Sensitivity analysis of associations between clinical phenotypes and third tertiles of dietary intakes derived from FFQ-based self-reported intake or metabolite score, adjusted for age, ethnicity and BMI (n=8,391), with first tertile (either intake or score) as reference. β-coefficients were obtained from independent linear regression (for HOMA-IR, FMI, cIMT) or logistic regression (for T2D, hypertension) for each food/beverage, based on the following sample sizes: T2D (n=1,040 T2D, n=7,351 non-T2D), hypertension (n=2,411 with hypertension, n=5,980 without hypertension). Association strength represents log-transformed *P* values. Circle border represents significant associations (*P*<0.05). HOMA-IR, homeostatic model assessment for insulin resistance; T2D, type 2 diabetes; FMI, fat mass index; cIMT, carotid intima-media thickness.

**Extended Data Fig. 5.**
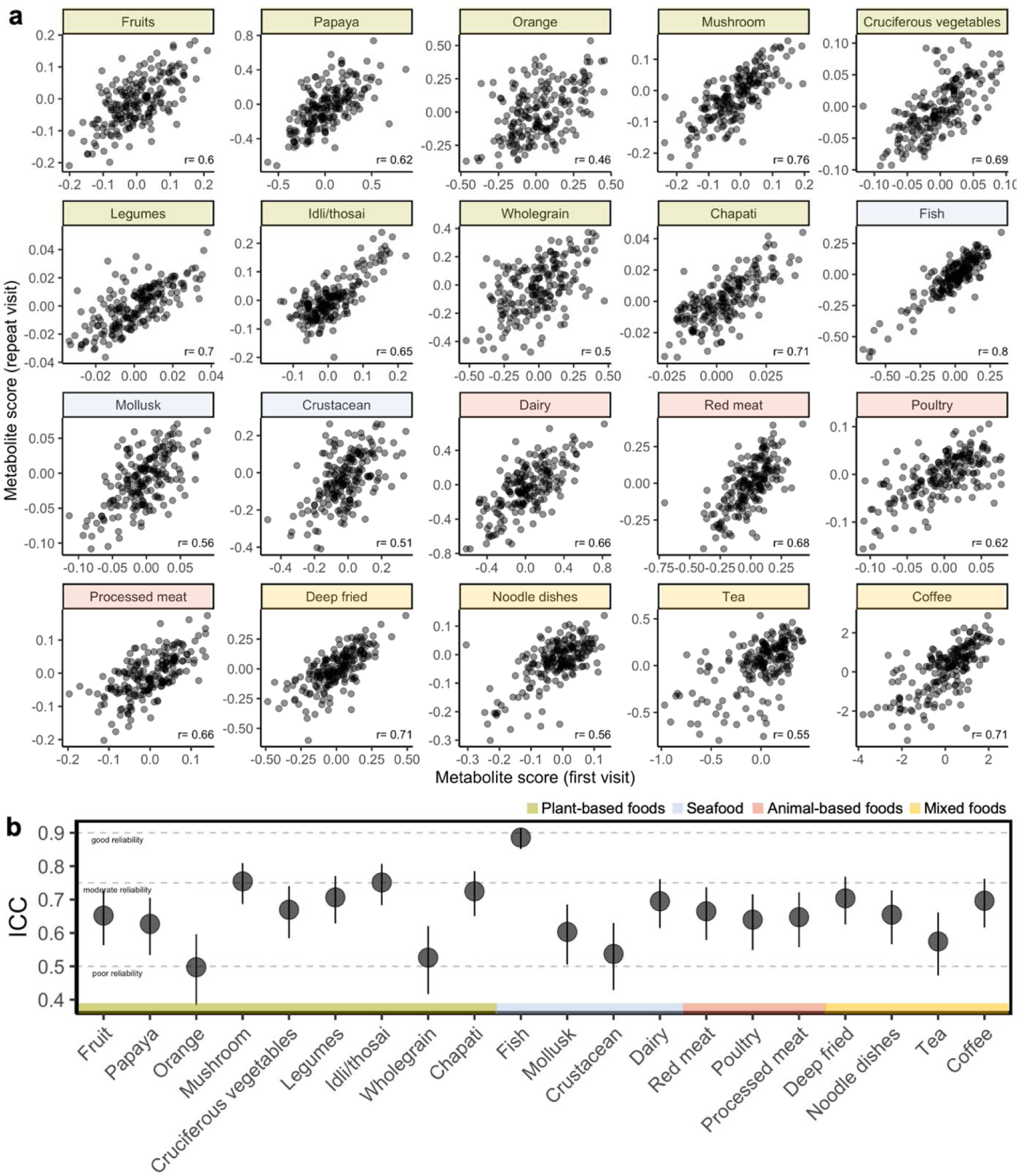
a) Scatter plots showing agreement (spearman *r*) between metabolite composite scores at first and repeat visits for each food and beverage in the repeat visit (n=195 pairs of individuals). b) Intraclass correlation coefficients (ICC) showing reliability of metabolite scores across first and repeat visits.

